# A Warm-start Digital CRISPR-based Method for the Quantitative Detection of Nucleic Acids

**DOI:** 10.1101/2021.06.10.21258725

**Authors:** Xiaolin Wu, Yie Hou Lee, Timothy K. Lu, Hanry Yu

## Abstract

Nucleic acids-based molecular diagnostic tools incorporating the CRISPR/Cas system are being developed as rapid and sensitive methods for pathogen detection. However, most CRISPR/Cas-based diagnostics lack quantitative detection ability. Here, we report Warm-Start RApid DIgital Crispr Approach (WS-RADICA), which uses commercially available digital chips for the rapid, sensitive, and quantitative detection of nucleic acids. WS-RADICA detected as little as 1 copy/μl SARS-CoV-2 RNA in 40 min (qualitative detection) or 60 min (quantitative detection). WS-RADICA can be easily adapted to various digital devices: two digital devices were evaluated for both DNA and RNA quantification, with linear dynamic ranges of 0.8-12777 copies/µL for DNA and 1.2-18391 copies/µL for RNA (both R^2^ values > 0.99). Moreover, WS-RADICA had greater sensitivity and inhibitor tolerance than a bulk RT-LAMP-Cas12b reaction and similar performance to RT-qPCR and RT-dPCR. Given its speed, sensitivity, quantification capability, and inhibitor tolerance, WS-RADICA shows great promise for a variety of applications requiring nucleic acid quantification.

## INTRODUCTION

Because they play important roles in nearly all biological processes, DNA and RNA are usually used as detection targets to indicate the presence of a biological entity. Technologies for nucleic acid quantification are needed in diverse areas, ranging from biomedical research to clinical diagnostics to environmental protection. The widely used RT-qPCR methods offer advantages in speed and sensitivity but require precise thermal cycling and high PCR efficiency. Sometimes accurate quantitative results cannot be acquired because of the PCR inhibitors present in the sample^1-3^. Digital PCR (dPCR) solves this problem by subdividing the PCR reaction into thousands of independent reactions and utilizing endpoint fluorescence levels in these partitions to determine amplification positive or negative compartments and calculate the target concentrations^4^. Since partially inhibited partitions with impaired PCR efficiencies still score positive for analysis, dPCR is more tolerant to inhibition than qPCR^2, 3, 5^. dPCR, however, requires a longer reaction time (more than 3h) because of the slow heat transfer rate among the partitions during thermal cycling. Many rapid isothermal amplification methods, such as recombinase polymerase amplification (RPA) or loop-mediated isothermal amplification (LAMP)^6, 7^, also detect nucleic acids with high sensitivity; however, these methods can suffer from nonspecific amplification and are often not quantitative^8, 9^.

Recently, CRISPR/Cas-based biosensing has been used alone or coupled with isothermal amplification for nucleic acid detection^10^. The trans-cleavage activity of Cas protein enables the degradation of synthetic nucleic acid reporters (quenched fluorescent reporters) when the target is complementary to the designed CRISPR RNA (crRNA) sequence, thus converting a nucleic acid sequence signal to a fluorescence signal. Methods such as SHERLOCK, HOLMES, and DETECTR were developed by coupling isothermal amplification and CRISPR/Cas biosensing to detect a variety of viruses as well as cancer gene targets^11-17^. Some one-pot methods, as well as amplification-free CRISPR/Cas methods, have also been used in combatting the SARS-CoV-2 pandemic^18-22^. Although they have high sensitivity, such methods are intended for qualitative detection without quantification. Yet, it would often be valuable to be able to quantify the results, which are needed to predict disease severity^23^.

To achieve rapid quantification, our group and three other groups recently have developed digital CRISPR-based methods for nucleic acid quantification^24-28^. Most of these methods were performed at 37ºC or 42ºC, with the sample preparation at low temperature to prevent target amplification or target cleavage before loading on the chip^24, 26-28^. Another method elevated the reaction temperature by coupling low-temperature reverse transcription dual-priming isothermal amplification (RT-DAMP) with Cas12a, which allows quantification at 52ºC but has low sensitivity. The additional step of adding the pyrophosphatase and phosphorothioated primers also adds more steps to these methods^25, 29^. Furthermore, these methods are limited to lab-made devices or self-developed image processing protocols, which restrict the application of digital CRISPR. Hence, there is still a need for warm-start rapid and sensitive methods for the quantification of nucleic acids that can be easily adapted to multiple kinds of digital devices.

Here, we report the development of a Warm-Start RApid DIgital Crispr Approach (WS-RADICA) for quantitative nucleic acid detection. In this method, digital detection is realized by dividing a one-pot warm-start RT-LAMP and CRISPR/Cas12b-based detection assay into thousands of nanoliter or subnanoliter reaction wells, and the quantitative result is obtained by counting the ratio of fluorescence-positive wells to all reaction wells. The warm-start activity of RT-LAMP prevents amplification at room temperature before sample partitioning and enables accurate digital quantification. A proof of concept assay was developed by targeting the nucleocapsid (N) gene of SARS-CoV-2. This method allowed qualitative detection in 40 min and quantitative results in 60 min in a 60 ºC water bath and could detect as little as 1 copy/uL of SARS-CoV-2 RNA. Moreover, we explored the applicability of this method in two different commercially available digital devices for the quantitative detection of SARS-CoV-2 RNA in a human genomic DNA background. The performances were comparable for WS-RADICA, RT-qPCR, and RT-dPCR reactions. WS-RADICA showed better sensitivity and inhibitor tolerance than the bulk RT-LAMP-Cas12b method. WS-RADICA demonstrates enhanced speed and sensitivity compared to other methods and tolerates inhibitors; therefore, it offers an attractive option for nucleic acid quantification.

## EXPERIMENTAL SECTION

### Materials

The sequences of primers, crRNA, and FQ reporters in this study, listed in Supplementary Table 1, were synthesised by Integrated DNA Technologies. Plasmids containing the N gene from each virus genomes (SARS-CoV-2, SARS-CoV, and MERS-CoV) were purchased from Integrated DNA Technologies. The synthetic RNA covering 99.9% of the bases of the SARS-CoV-2 viral genome was purchased from Twist Bioscience (Genbank ID: MN908947.3). The DNA and RNA concentrations were measured by dPCR or RT-dPCR assay.

### Bulk RT-LAMP-Cas12b reactions

The DNA/RNA target samples were mixed with 1.6 µM FIP primers, 1.6 µM BIP primers, 0.2 µM F3 primers, 0.2 µM B3 primers, 0.4 µM LoopF primers, 0.4 µM LoopB primers, 1.4 mM dNTPs, 8 mM MgSO_4_, 2 µM FQ-5T reporters, 0.96 U/µL Bst 2.0 WarmStart polymerase (New England Biolabs), 0.3 U/µL WarmStart RTx Reverse Transcriptase (only used for RNA detection, New England Biolabs), 1 U/µL RNase Inhibitor (New England Biolabs), 50 mM Taurine, 50 nM Cas12b (Magigen Biotechnology), and 50 nM crRNA in 1x isothermal amplification buffer (New England Biolabs), unless otherwise indicated. To detect a LAMP signal, 250 mM SYTO-82 fluorescent nucleic acid stain was added to the reaction. The reaction mixture was incubated at 60°C and fluorescence kinetics were monitored for 1-2 h. To mimic the complexity of clinical samples, 1 ng/µL human genomic DNA (Roche) was also added to the reactions.

### WS-RADICA reactions by Clarity digital chip

The WS-RADICA reaction was prepared by adding 1x Clarity™ JN solution (JN Medsys) to the RT-LAMP-Cas12b bulk reactions described above and partitioned on a Clarity digital chip^30^. 15 µL reaction mixtures were loaded onto the digital chip using Clarity™ autoloader, followed by treatment with the Clarity™ sealing enhancer and sealing with 230 μL Clarity™ sealing fluid. The tube containing the digital chip was warmed in a water bath at 60°C for 1 hour, unless otherwise indicated. After incubation, the end-point fluorescence in the 10,000 partitions was detected by a Clarity™ Reader, and positive partition percentages, as well as input nucleic acids copy number, were calculated by Clarity™ software.

### WS-RADICA reactions by QIAcuity digital nanoplate

The WS-RADICA reaction was prepared by adding 1x reference dye to the RT-LAMP-Cas12b bulk reactions stated above and partitioned on QIAcuity digital nanoplates. 40 µL reaction mixtures were loaded onto the QIAcuity digital nanoplate and into the QIAcuity Digital PCR System. In the QIAcuity Digital PCR machine, the reactions were automatically partitioned into 26,000 microwells followed by 60°C incubation for 1 hour and end-point fluorescence detection. Positive partition percentages, as well as input nucleic acid copy number, were calculated by QIAcuity software.

### RT-dPCR reaction

The USCDC N2 assay for SARS-CoV-2 detection was purchased from Integrated DNA Technologies. Serial dilutions of RNA targets were mixed with 1x USCDC N2 assay, 1x TaqMan™ Fast Virus 1-Step Master Mix (Applied Biosystems) and loaded on Roche Light Cycler. The reactions were incubated at 55°C for 5 min followed by 95°C for 20 s (one cycle), 95°C 10 s and 60°C 30 s (45 cycles). Fluorescent signals were monitored at the 60°C step, and Cq values (also called Ct values) were calculated automatically using Roche Light Cycler software.

### dPCR and RT-dPCR reaction

Plasmid DNA quantification by dPCR: Plasmids containing the N genes of SARS-CoV-2, SARS-CoV, and MERS-CoV were linearized with FastDigest ScaI (Thermo Scientific) then mixed with 1x USCDC N2 assay, 1x TaqMan™ Fast Advanced Master Mix (Applied Biosystems) and 1x Clarity™ JN solution (JN Medsys). The 15 µL reactions were loaded onto the Clarity digital chip using the method mentioned above, and the tubes containing the digital chip were transferred to a PCR machine using the following parameters (ramp rate = 1 °C/s): 95 °C for 15 min (one cycle), 95°C for 50s and 58°C for 90s (40 cycles), and 70°C for 5 min. The endpoint fluorescence of the partitions was detected with a Clarity™ Reader, and the input DNA copy numbers were calculated by Clarity™ software.

RNA detection by RT-dPCR: Serial dilutions of RNA targets were mixed with 1x USCDC N2 assay, 1x TaqMan™ Fast Virus 1-Step Master Mix (Applied Biosystems) and 1x Clarity™ JN solution (JN Medsys) followed by incubation at 55°C 5 min. After incubation, the reactions were partitioned on the Clarity digital chip and transferred to a PCR thermocycler with the following parameters (ramp rate = 1 °C/s): 95°C for 15 min (one cycle), 95°C for 50s and 58°C for 90s (40 cycles), and 70°C for 5 min. The endpoint fluorescence of the partitions was detected using a Clarity™ Reader and the input RNA copy numbers were calculated by Clarity™ software.

### Inhibitor tolerance test of the reactions

To test the inhibitor tolerance, 2.5 U/mL heparin, 0.01% Sodium dodecyl sulphate (SDS), 1 mM ethylenediaminetetraacetic acid (EDTA), or 5% ethanol was added to the reactions mentioned above. For bulk reactions (RT-qPCR and bulk RT-LAMP-Cas12b), the 1h end-point fluorescent signals of reactions with inhibitors were divided by the reactions without inhibitors, and the corresponding ratio was taken as the percentage of efficiency. For digital reactions (RT-dPCR and WS-RADICA), the positive partition percentage of the reactions with inhibitors was divided by that of the reactions without inhibitors, and the corresponding ratio was taken as the percentage of efficiency.

## RESULTS AND DISCUSSION

### Principle of WS-RADICA

WS-RADICA combines warm-start RT-LAMP and Cas12b in a one-pot reaction and digitalizes the reaction into thousands of nanoliter or subnanoliter reactions for quantitative results. The principle of WS-RADICA is illustrated in Figure 1. First, the DNA or RNA is extracted from its source and assembled with WarmStart RTx reverse transcriptase, *Bst* 2.0 WarmStart DNA polymerase, LAMP primers, Cas12b/crRNA, and a quenched fluorescent (FQ) reporter. Then the reaction mixture is divided into thousands of partitions on a digital chip, followed by incubating in a 60ºC water bath. In each of the independent partitions containing the target, the RT-LAMP reaction starts to amplify the DNA/RNA exponentially. Simultaneously, the amplified DNA is identified by Cas12b/crRNA by sequence complementarity, which in turn triggers Cas12b, via its trans cleavage activity, to cut the FQ reporter and generate increased fluorescence (Figure 1B). Thus the partitions containing target DNA or RNA emit a positive fluorescent signal while the partitions without target DNA or RNA emit only the baseline signal (background).

**Fig. 1.**
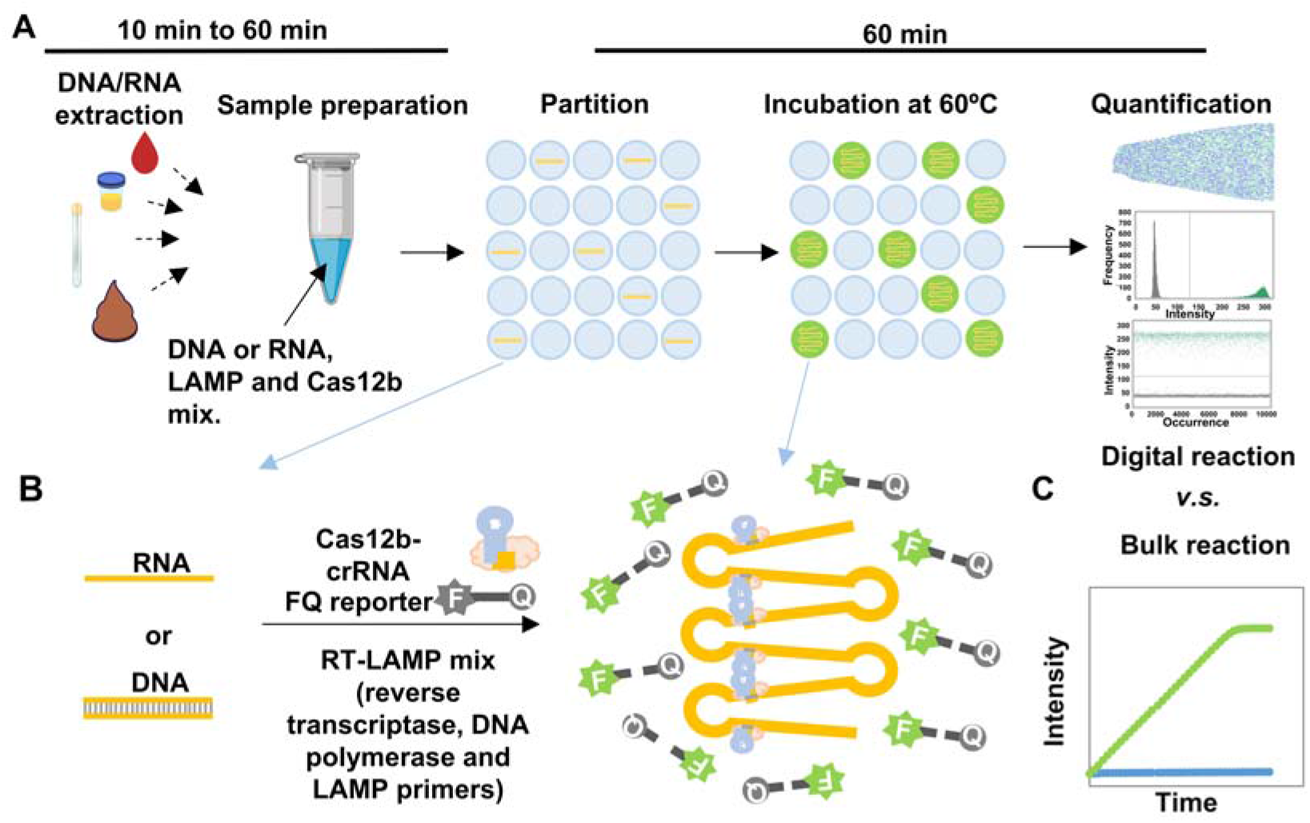
Schematic illustration of WS-RADICA. (A) Overview of the WS-RADICA process. Nucleic acids (DNA and RNA) are extracted from different types of samples, then mixed with RT-LAMP reagent and Cas12b/crRNA/FQ reporter mix. The reaction mixtures can be subdivided into thousands of partitions by digital chips, followed by incubation at 60ºC for 1 h. The partitions containing the target yield a much higher fluorescent signal than the partitions without targets, and the end-point results are detected by a fluorescent detector to calculate the proportions of positive partitions. (B) Reactions in the positive partitions. The DNA/RNA, RT-LAMP reagents, and Cas12b/crRNA/FQ reporter are mixed in a one-pot format in each partition. The target DNA/RNA can be amplified by RT-LAMP mixtures into looped structures. Because the amplified targets are complementary with crRNA, they bind to a Cas12b/crRNA complex, triggering the trans cleavage of Cas12b protein to cut the FQ reporters, which in turn results in a fluorescent signal. (C) Schematic concept of the bulk RT-LAMP-Cas12b assay.

As the RT-LAMP reaction is warm-start, the reaction will be inhibited at temperatures below 45ºC and will start only after the samples are partitioned and incubated at 60ºC, which enables room-temperature reaction setup and increases the accuracy and consistency of the results. The Cas enzyme we used was the thermostable Cas12b from *Alicyclobacillus acidiphilus* (AapCas12b), which has been shown to be compatible with the one-pot RT-LAMP reaction with high sensitivity^19^. Compared to a bulk reaction (Figure 1C), the digital reaction is quantified by assigning each positive and negative partition as “one” or “zero” and calculating the percentage of positive partitions. As the partitions are physically separated, the digital reaction eliminates interference among individual reaction wells.

### Optimization of the RT-LAMP-Cas12b assay for digital detection

Systematic studies of one-pot RT-LAMP-Cas12b reaction kinetics were conducted to optimize the assay performance. To increase the signal-to-noise ratio (S/N) in each partition and reduce the reaction time, the FQ reporter with 5 thymine (T) was chosen because it showed the highest S/N and reaction rate compared to those of other FQ reporters with different base compositions (poly-A, poly-T, poly C, poly G and poly AT, Figures 2A and S1) or different lengths (5-20 nucleotides, Figures 2B and S2) in bulk reaction. Taurine was also added to the reaction as it improved the reaction kinetics (Figures 2C and S3), which is consistent with a previous study^19^. Also, the concentrations of the enzymes in this reaction were optimized to get the highest positive partition ratio in the presence of the same concentrations of RNA, and the resulting optimal concentrations were 0.3 U/µL WarmStart RTx reverse transcriptase (Figures 2D and S4), 0.96 U/µL *Bst* 2.0 WarmStart DNA polymerase (Figures 2E and S5), and 50 nM Cas12b/crRNA (Figure 2F).

**Fig. 2.**
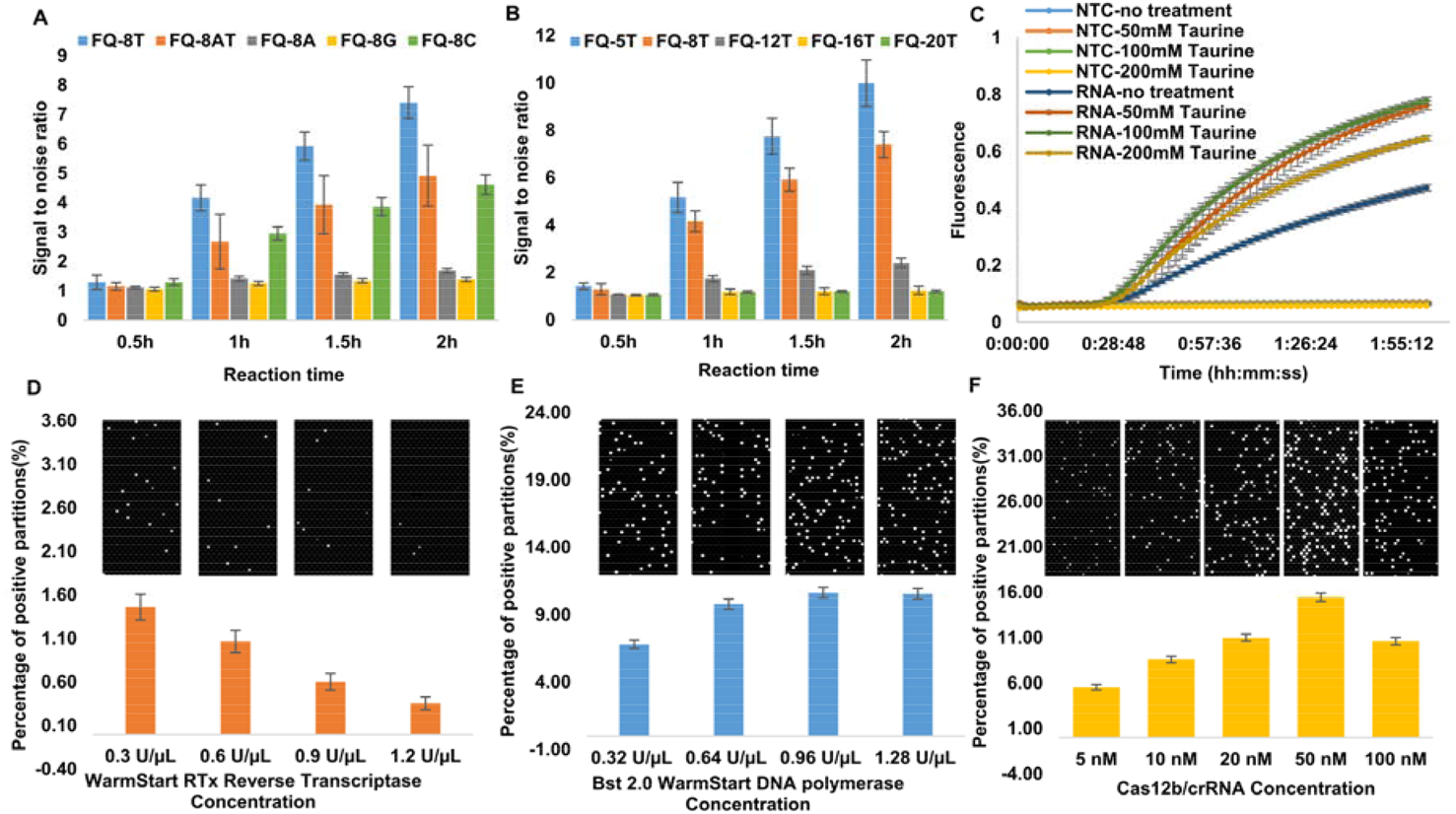
Optimization of WS-RADICA. (A) Comparison of bulk RT-LAMP-Cas12b reaction under FQ reporter with different base compositions (poly A, poly T, poly C, poly G, and poly AT). (B) Comparison of bulk RT-LAMP-Cas12b reaction with different lengths of FQ reporter (5 nt to 20 nt). (C) The effect of taurine on the bulk RT-LAMP-Cas12b reaction. (D) Digital RT-LAMP-Cas12b reactions with several concentrations of reverse transcriptase. (E) Digital RT-LAMP-Cas12b reactions with several concentrations of polymerase. (F) Digital RT-LAMP-Cas12b reactions with several concentrations of Cas12b/crRNA. For figures D, E and F, the upper panels are representative end-point fluorescence images while the lower panels show the proportion of positive partitions in a total number of around 26,000 partitions for each replicate.

Using the optimized parameters above, we subsequently tested the real-time performance of bulk reactions at concentrations of RNA ranging from 1 to 18391 copies/µL. In the one-pot assay, RT-LAMP reactions were monitored using a SYTO-82 orange fluorescent nucleic acid stain (Figure 3A), and Cas12b reactions were monitored using a green FQ reporter (Figure 3B). From the result, the RT-LAMP-Cas12b can detect as little as 6 copies/µL RNA in the bulk reaction, while 1 copy/µL RNA showed no difference to non-targeted control (NTC). From the orange fluorescent signal (Figure 3A), the exponential amplification of the target started at around 9-15 min and reached a plateau at 25-35 min; the time needed to reach the plateau correlated with the concentration of target RNA. At the time of around 47 min, the NTC reaction started to have non-specific amplification signals, which are common in LAMP due to the interaction of multiple primers^9^. Non-specific amplification is hard to avoid with LAMP because six primers are involved; thus, an additional CRISPR/Cas-based detection is vital to the specificity of this assay. From the green fluorescent signal (Figure 3B), the trans cleavage activity of Cas12b started to be detected 6 min later than LAMP activity, suggesting that a short time is needed for the amplified DNA to accumulate and for Cas12b to detect it. No increased green fluorescent signal was detected in the NTC sample, which is different from the LAMP signal and indicates the specificity of the Cas12b reaction. Also, the Cas12b was still proceeding without reaching a plateau even after 2 h of reaction time, indicating that the end-point fluorescence of the bulk reaction may be used as a semi-quantitative marker. These results confirmed the short reaction time of the one-pot RT-LAMP-Cas12b reaction and the specificity of the reaction obtained by adding Cas12b/crRNA.

**Fig. 3.**
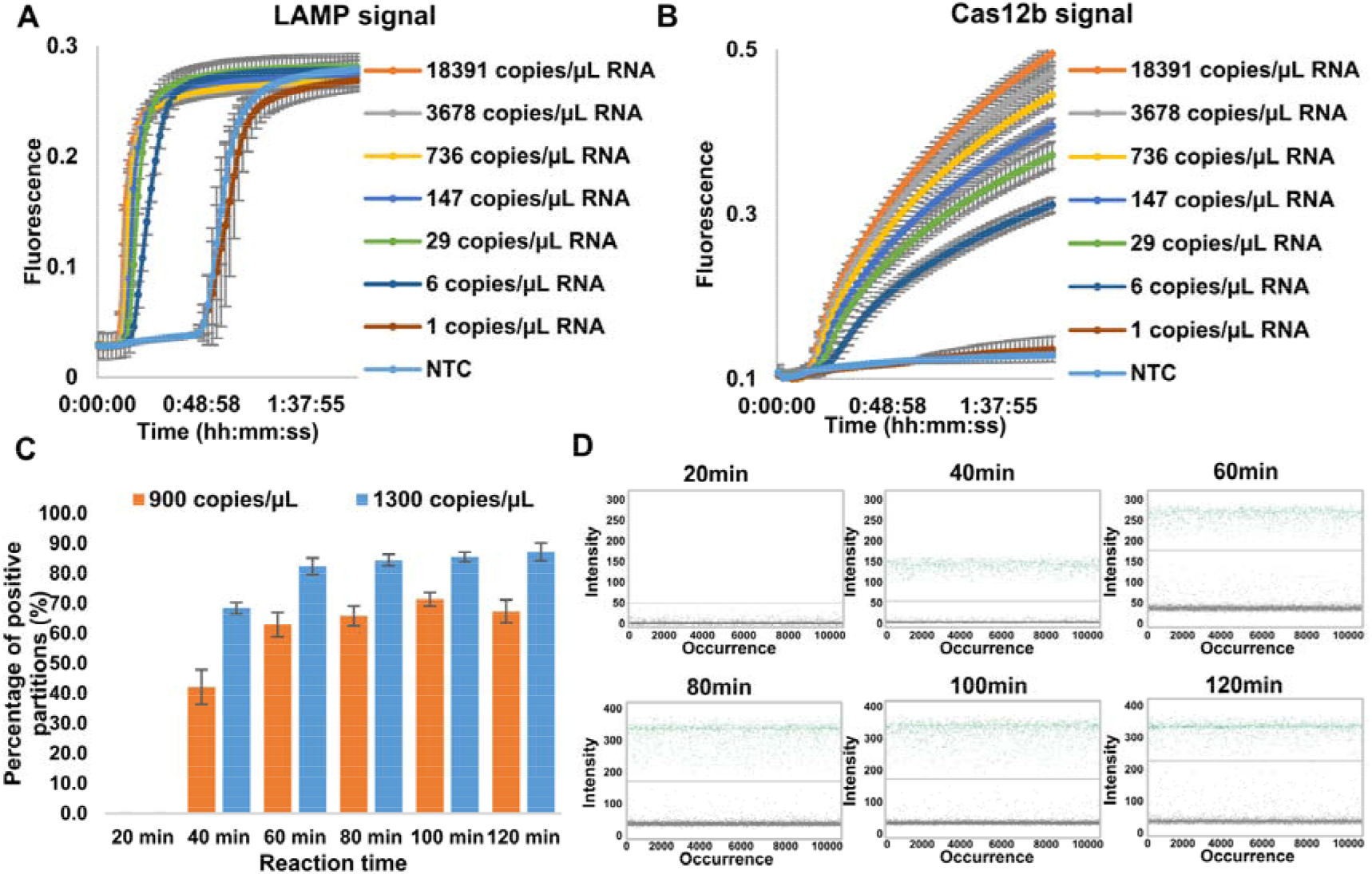
Time-course reactions of WS-RADICA. (A, B) Real-time reactions of bulk RT-LAMP-Cas12b with a range of concentrations of RNA or non-targeted control. The LAMP amplified signal was detected by SYTO82 fluorescent nucleic acid stain (which exhibits orange fluorescence upon binding to nucleic acids, A) and Cas12b signals by FQ reporter (which exhibits green fluorescence upon cutting by Cas12b, B). (C) Comparison of positive partition percentages of WS-RADICA with different reaction times at 60ºC for several concentrations of targets. (D) End-point fluorescence results of the same WS-RADICA at incubation times of 20, 40, 60, 80, 100, and 120 min, at 60ºC.

We then investigated the reaction time of the reaction on a digital device by measuring the percentages of positive partitions at different reaction times (Figures 3C and 3D). From the time-course result (Figure 3C), most of the positive partitions could be detected at 40 min for the targets at different concentrations, indicating that 40 min was enough for qualitative detection, which is consistent with the bulk reaction. A slight difference from the bulk reaction was that some partitions containing the targets could not be distinguished from the background and were assigned as negative by the software due to the slightly low fluorescence level at 40min. As time went on, the fluorescent signals increased in these partitions and the percentage of positive partitions plateaued at around 60 min, suggesting that all the partitions containing the target could be distinguished from negative partitions at 60 min. After 60 min, the reaction was still proceeding and the fluorescent signals in the positive partitions were still increasing (Figure 3D), while the ratio of positive partitions remained constant (Figure 3C). These results indicate that 40 min incubation is enough for qualitative detection and 60 min incubation is enough for quantitative detection by WS-RADICA.

### Performance of WS-RADICA and its applications on different digital devices

To investigate the performance of WS-RADICA, the specificity, quantification capability, and applicability of this method were evaluated. DNA or RNA containing the SARS-CoV-2 sequence was added to the reaction as the target and LAMP primers and crRNA targeting N gene of SARS-CoV-2 were used to evaluate the performance of the digital reaction on a commercially available Clarity digital chip ^19, 30^. First, the specificity of WS-RADICA was tested by comparing the results of SARS-CoV-2 DNA with SARS-CoV DNA, MERS-CoV DNA, and a human genomic DNA control. From the result (Figure 4A), positive partitions were detected only in the target sample (SARS-CoV-2 DNA) but not in the controls (SARS-CoV DNA, MERS-CoV DNA, and a human genomic DNA control), indicating the high specificity of this method in distinguishing similar sequences.

**Fig. 4.**
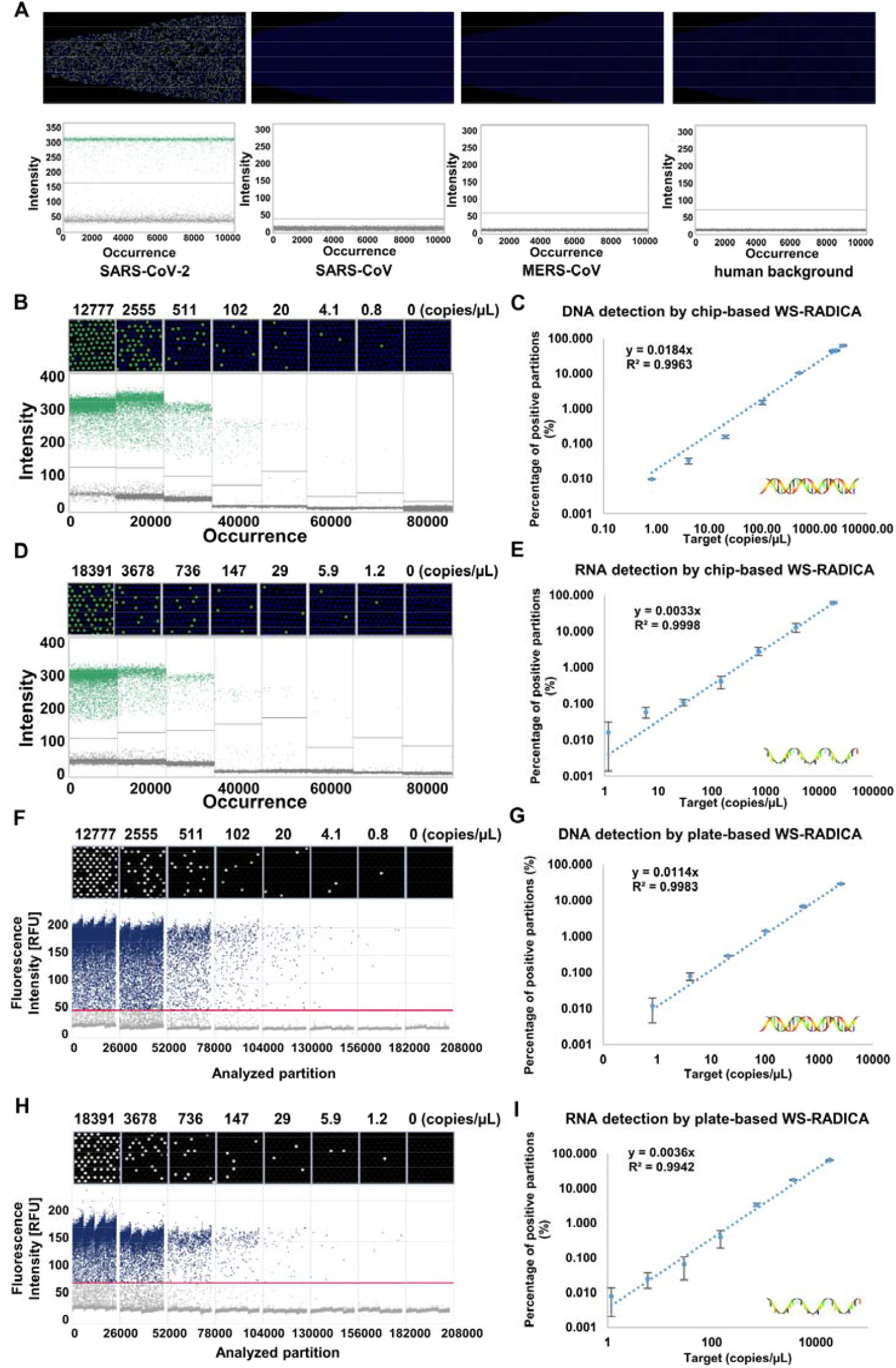
Quantitative detection of nucleic acids by WS-RADICA on different digital detection platforms. (A) Specificity of WS-RADICA reaction. Primers and crRNA targeting SARS-CoV-2 were used to test the WS-RADICA reaction for SARS-CoV-2, SARS-CoV, and MERS-CoV sequences with a human DNA background. The upper panels are representative end-point fluorescence images while the lower panels show the scatter plot representing a total number of around 10,000 partitions for one sample. (B, C) WS-RADICA with different concentrations of DNA target using a Clarity digital chip. (D, E) WS-RADICA with different concentrations of RNA target using a Clarity digital chip. (F, G) WS-RADICA with different concentrations of DNA target using QIAcuity digital nanoplate. (H, I) WS-RADICA with different concentrations of RNA target using QIAcuity digital nanoplate. For figures B, D, F, and H, the upper panels are representative end-point fluorescence images while the lower panels show the scatter plot representing a total number of around 10,000 partitions (B, D) or 26,000 partitions (F, H) for one sample. Figures C, E, G and I represent the correlation and linear relationship between the input target concentrations and positive partition percentages. Each concentration consists of at least three replicates.

As it would be useful to be able to quantify nucleic acids with this method as well as simply to detect their presence, WS-RADICA was evaluated for its ability to distinguish different concentrations of SARS-CoV-2 DNA (5 serial dilutions ranging from 12777 to 0.8 copies/µL) or RNA (5 serial dilutions ranging from 18391 to 1.2copies/µL). From the scatter plot results of about 10,000 partitions (Figures 4B and 4D), positive partitions decreased in more diluted samples and no positive partitions were detected in the NTC sample. Also, there was a strong linear relationship (R^2^ = 0.99) between the target concentrations and the percentage of positive partitions in both DNA and RNA samples (Figures 4C and 4E). The measured concentration of WS-RADICA based on Poisson distribution was lower than the input target concentration, which can possibly be attributed to “molecular dropout” or low filling rate of the microwells, as has been previously reported for dPCR, dRPA, and dLAMP reactions^3, 24, 25, 31^. Nevertheless, WS-RADICA has a single reaction temperature and faster reaction time, which are important advantages. Also, the reactions noted above were observed in the presence of 1 ng/µL human DNA, indicating that WS-RADICA’s quantification capability was not affected by background nucleic acids.

Having shown the specificity and quantification capability on the commercially available Clarity digital chip, we asked whether WS-RADICA could be easily used with other digital devices which had been previously designed for digital PCR and which are in common use. A plate-based QIAcuity Digital PCR System was tested for this purpose. The same one-pot reactions were digitalized using the sample partition system on QIAcuity to divide the samples into 26,000 individual wells in the plate followed by 60ºC incubation for 60min. For both DNA (0.8-12777 copies/µL) and RNA (1.2-18391 copies/µL), the number of positive wells in the plate increased when more targets were added to the reaction (Figures 4F and 4H). A strong linear relationship (R^2^ = 0.99) was also found between the target concentration and the positive well ratio (Figures 4G and 4I). Although the relationship between the target concentration and positive partition percentage was slightly different for the two digital devices due to their different partition volumes, accurate quantification results were obtained based on the standard curve from the same device. Therefore, WS-RADICA can be easily adapted to different digital devices, offering a faster alternative for those who already have digital PCR machines for nucleic acid quantification.

### Comparison of WS-RADICA with other detection methods

To determine whether WS-RADICA is competitive with other nucleic acid detection methods, such as RT-qPCR, RT-dPCR, and RT-LAMP-Cas12b bulk reaction, we performed these four methods and compared their detection of the same concentrations of SARS-CoV-2 RNA (from 18391 copies/µL to 1 copy/µL) in the presence of a human genomic DNA (1 ng/µL) background. For the PCR-based experiments (RT-qPCR and RT-dPCR), the FDA EUA-approved CDC assay targeting the SARS-CoV-2 N gene was used; and for the CRISPR/Cas based methods (RT-LAMP-Cas12b bulk reaction and WS-RADICA), primers and crRNA targeting the same N gene region of SARS-CoV-2 were used^19^. For each RNA dilution, more than three replicates were tested. For RT-qPCR, the serial dilutions of SARS-CoV-2 RNA gave threshold cycle numbers (Ct) ranging from 17.8 to 32.2, and there was a very strong linear relationship between the log of target concentration and Ct, as well as sensitivity corresponding to the detection of 1 copy/µL (Figures 5A and S6). For RT-dPCR, the RNA concentration (18391 copies/µL) was far above the upper linear range and resulted in more than 99.6% of the partitions being positive. A good linear relationship between the target concentration and measured concentration was also observed for the RT-dPCR results, corresponding to the detection of a large range of concentrations, from 3678 copies/µL to 1 copy/µL of RNA (Figures 5B and S7). However, for RT-dPCR the reaction took much longer (more than 3 h), which may hinder its widespread use.

**Fig. 5.**
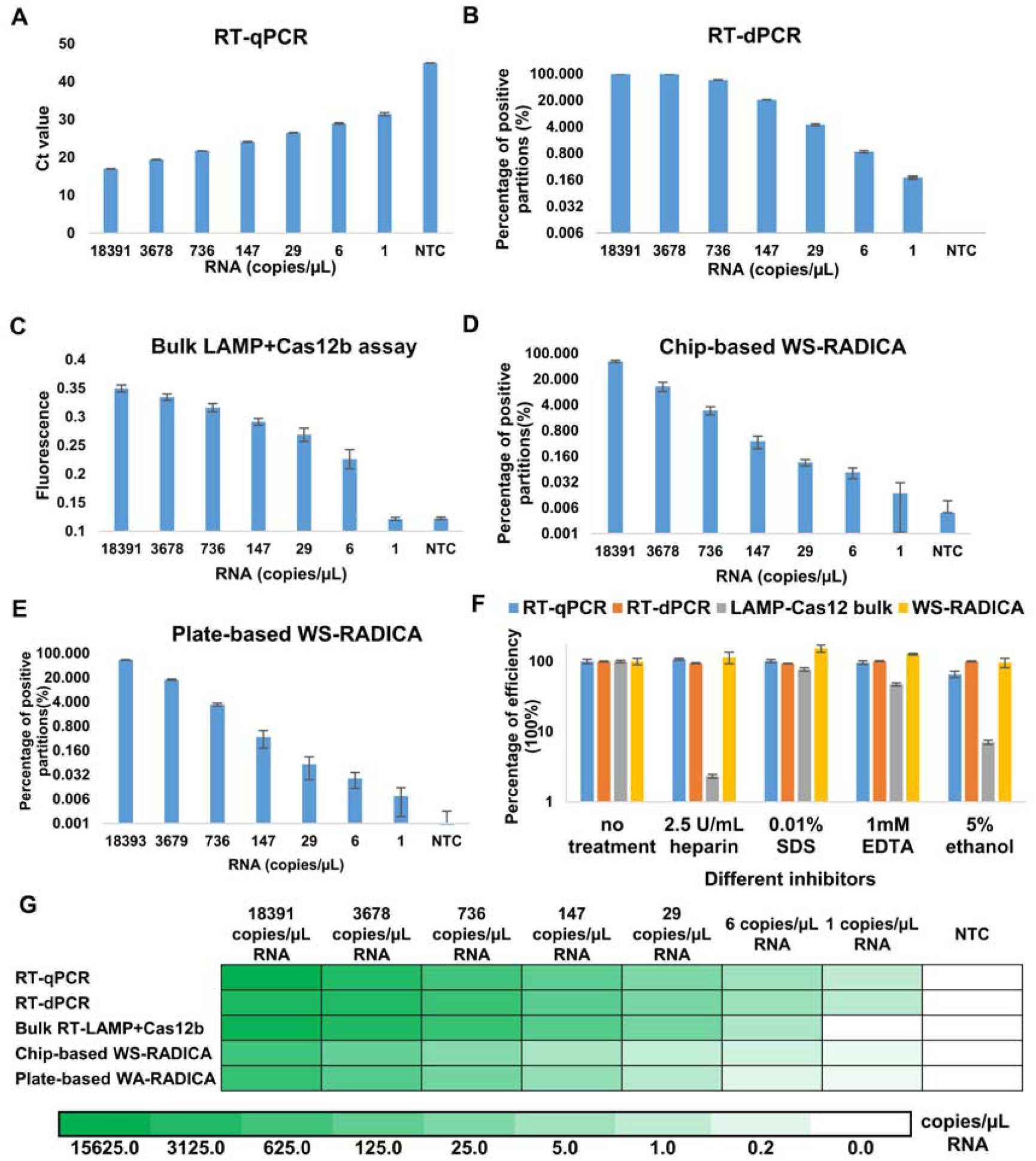
Comparison of WS-RADICA with other detection methods. The detection sensitivity of RT-qPCR (A), RT-dPCR (B), Bulk RT-LAMP-Cas12b assay (C), WS-RADICA by Clarity digital chip (D), and WS-RADICA by QIAcuity digital nanoplate (E) using different concentrations of SARS-CoV-2 RNA as target with the background of 1 ng/µL human genomic DNA. (F) Comparison of the effects of reaction inhibitors on RT-qPCR, RT-dPCR, Bulk RT-LAMP-Cas12b assay, and WS-RADICA. (G) Quantitative results of RT-qPCR, RT-dPCR, Bulk RT-LAMP-Cas12b assay, WS-RADICA by Clarity digital chip and WS-RADICA by QIAcuity digital nanoplate in detecting SARS-CoV-2 RNA at various concentrations. Heat map represents the measured concentrations of each method using the corresponding quantitative methods. Bulk reactions (RT-qPCR and bulk RT-LAMP-Cas12b) results were quantified using corresponding standard curves. Digital reaction results obtained with various methods (RT-dPCR, WS-RADICA by Clarity digital chip, and WS-RADICA by QIAcuity digital nanoplate) were quantified by Poisson distribution.

Compared to PCR-based methods, the RT-LAMP-Cas12b-based method is relatively simple, as the isothermal amplification does not require thermal cyclers. As to sensitivity, the bulk reaction was slightly weaker in detecting low copy number samples, resulting in a detection sensitivity of 6 copies/µL RNA (Figure 5C). Although end-point fluorescence of the bulk reaction can be used as a semi-quantitative marker, the relatively weak linear relationship between signal and target suggests that the RT-LAMP-Cas12b bulk reaction is not suitable for quantification (Figure S8). Because with WS-RADICA the whole reaction is divided into thousands of independent reactions, calculating the percentage of positive partitions enables quantification. Also, the sensitivity of the reaction increased by taking the benefit of the confinement effect on local concentration^26^. One molecule confined in a 1 nL well is equal to 1.66 fM local concentration, which is also equal to 10,000 molecules in a 10 µL bulk reaction. Thus, we still see positive partitions in WS-RADICA at 1 copy/µL but not in the bulk reaction (Figures 5D and 5E). Moreover, WS-RADICA widened the linear dynamic range for quantitative detection beyond that achieved with RT-dPCR and enabled the quantification of 18391 copies/µL RNA on the same digital device. Overall, WS-RADICA has better sensitivity and quantitation ability than the bulk reaction and higher speed and wider dynamic range compared to RT-dPCR, which make it a promising alternative for nucleic acid quantification.

As inhibitors in the samples are likely to alter the reaction and might affect the accuracy of the result, we tested the effect of various inhibitors on the four detection methods used in the quantification experiments (Figure 5F). Heparin, normally used as an anticoagulant in blood, serum, or plasma samples, was found in a previous study to act on DNA polymerase and thus inhibit the reaction^1^. In our study, 2.5 U/mL heparin had no effect on RT-qPCR or RT-dPCR but greatly inhibited the bulk RT-LAMP-Cas12b reaction. In contrast, WS-RADICA tolerated heparin, showing similar quantitative results in the reactions with or without heparin. SDS, an ionic detergent commonly used in sample lysis, was also reported as a DNA polymerase inhibitor^32^. SDS slightly inhibited the bulk RT-LAMP-Cas12b reaction, whereas WS-RADICA recovered the reaction. Similarly, EDTA, which chelates metal ions, such as Mg^2+^, was found to inhibit the bulk RT-LAMP-Cas12b reaction but did not inhibit WS-RADICA. Also, ethanol, which is always used in nucleic acid purification, was tested for its effects on the four different methods. Both bulk RT-qPCR and the bulk RT-LAMP-Cas12b reaction were inhibited by ethanol to some degree. In contrast, the digital reactions (RT-dPCR and WS-RADICA) were more tolerant of ethanol. In accordance with previous studies on digital PCR^1, 3^, our experiments showed that digital reactions, not only RT-dPCR but also WS-RADICA, are more robust and less susceptible to inhibitors than bulk reactions, possibly because the individual micro-reactions alleviate the effect of inhibitors as long as the fluorescent signal can be differentiated from background in the presence of inhibitory substances^1, 5^. Thus, the digital detection format of the CRISPR reaction offers great advantages not only in sensitivity but also in inhibitor tolerance.

## CONCLUSIONS

In this study, we have developed the first universal warm-start digital CRISPR method and applied it to different types of digital detection machines. This method, WS-RADICA, combines the sensitivity of RT-LAMP, the specificity of Cas12b, and the absolute quantification ability of digital detection; these features enable the quantitative detection of nucleic acids in 1 hour. Compared to previous digital methods, the warm-start format of WS-RADICA allows the reaction to be prepared at room temperature, and digitalization is not restricted to a particular digital format^24, 26, 27^. Using WS-RADICA, a concentration of nucleic acid as low as 1 copy/µL could be quantitatively detected in two different digital formats. The results are specific and not influenced by similar sequences or human nucleic acid backgrounds. Comparable performances were achieved when WS-RADICA was compared to conventional RT-qPCR and RT-dPCR. We also demonstrated that, by digitalizing the bulk reaction, the sensitivity and the inhibitor resistance of the reaction greatly increased, which makes WS-RADICA a powerful tool for the absolute quantification of nucleic acids in a wide variety of samples.

As we showed by using SARS-CoV-2 as an example (Figures 4 and 5), WS-RADICA can be easily extended to a variety of clinical, research, and biomanufacturing applications, such as liquid biopsy, rare mutation detection, environmental monitoring, cancer research, and cell therapies^33-37^. Faster, more sensitive or amplification-free detection could be achieved with WS-RADICA using a more sensitive Cas enzyme, crRNA, FQ reporters, or a much smaller partition size^20, 22, 26, 38-40^. Crude samples, without the initial step of nucleic acid extraction, could also be used for single-cell detection^41^. Multiplexed methods could also be improved by utilizing different orthogonal Cas proteins or a spatially separated chip design^12, 16, 42, 43^. As WS-RADICA only requires one temperature for the reaction, it can be integrated with a portable heater and smartphone-based fluorescence detection for point-of-care quantification applications^44, 45^. Based on the superior performance in sensitivity, speed, inhibitor resistance, and quantitative detection and the great potential for improvement and applicability, we anticipate that WS-RADICA will be a promising quantitative molecular tool applicable to the clinical setting, as well as to research, biomanufacturing, and environmental and food industries.

## Data Availability

The authors declare that all data supporting the findings of this study are available within the paper (and its supplementary information files). Correspondence and requests for materials should be addressed to H.Y. and T.K.L.  

## ACKNOWLEDGMENTS

We thank Karen Pepper (MIT) for her careful editing and helpful comments on the manuscript. This work was supported by the National Research Foundation, Prime Minister’s Office, Singapore under its Campus for Research Excellence and Technological Enterprise (CREATE) programme, through Singapore MIT Alliance for Research and Technology (SMART): Critical Analytics for Manufacturing Personalised-Medicine (CAMP) Inter-Disciplinary Research Group.

## AUTHOR CONTRIBUTIONS

X.W., T.K.L., and H.Y. designed the research. X.W. developed the WS-RADICA method, performed the experiments, and analyzed the data. Y.H.L., T.K.L., and H.Y. provided mentorship and feedback. X.W. wrote the original draft and all authors reviewed and edited the manuscript.

## SUPPLEMENTARY DATA

Supplementary data related to this article are available online.

