## Supplementary data for "A Warm-start Digital CRISPR-based Method for the Quantitative Detection of Nucleic Acids"

**Items included in the supplementary data:**

**Supplementary Figures 1 to 8**

**Supplementary Tables 1 to 2**

**Supplementary References**

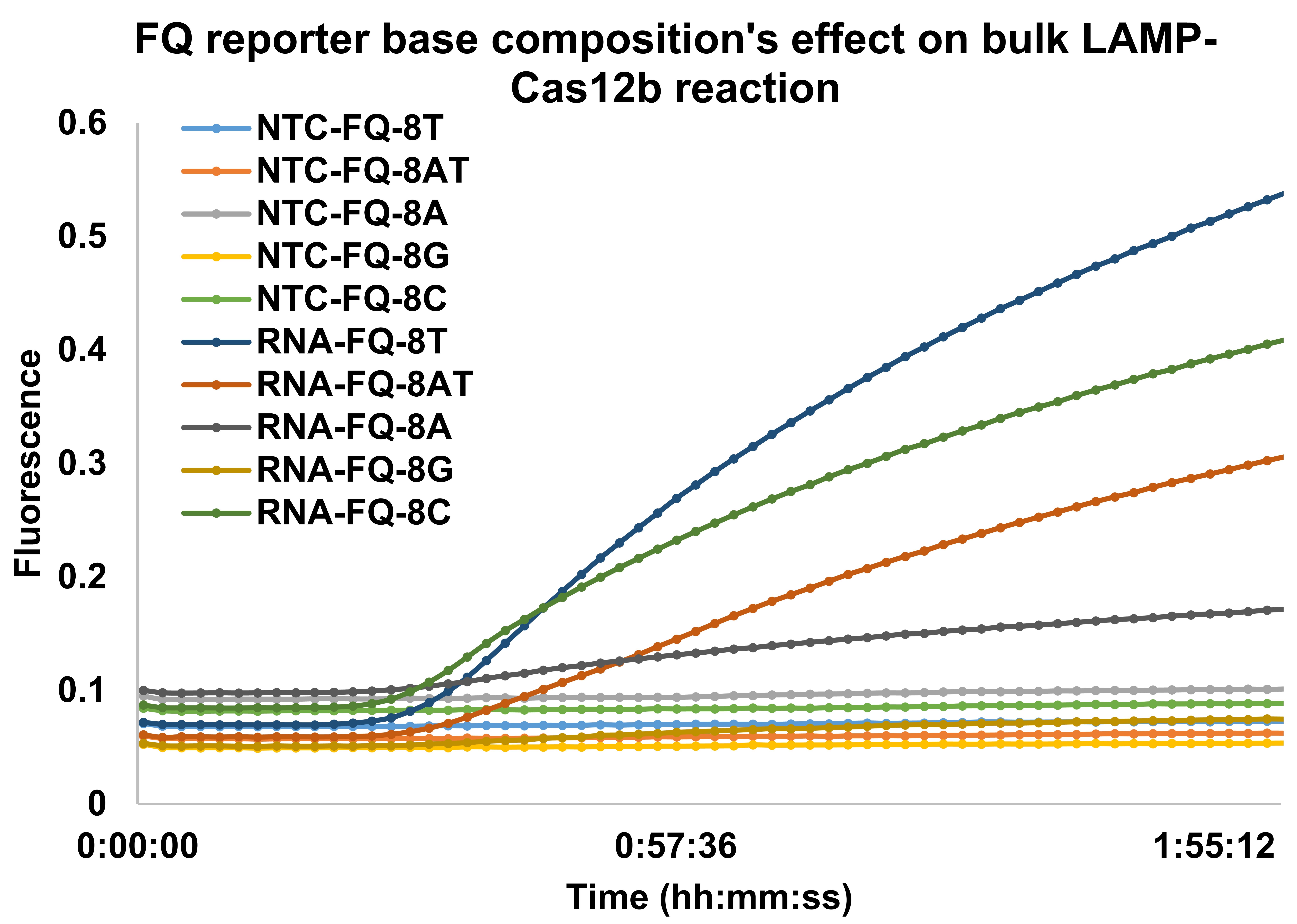

**Supplementary Fig. 1. FQ reporter base composition's effect on bulk RT-LAMP-Cas12b reaction.** Bulk RT-LAMP-Cas12b reactions with different FQ reporter sequences at the same length were monitored at 60 ºC. Fluorescent signals of reactions with target RNA and non-template control were compared.

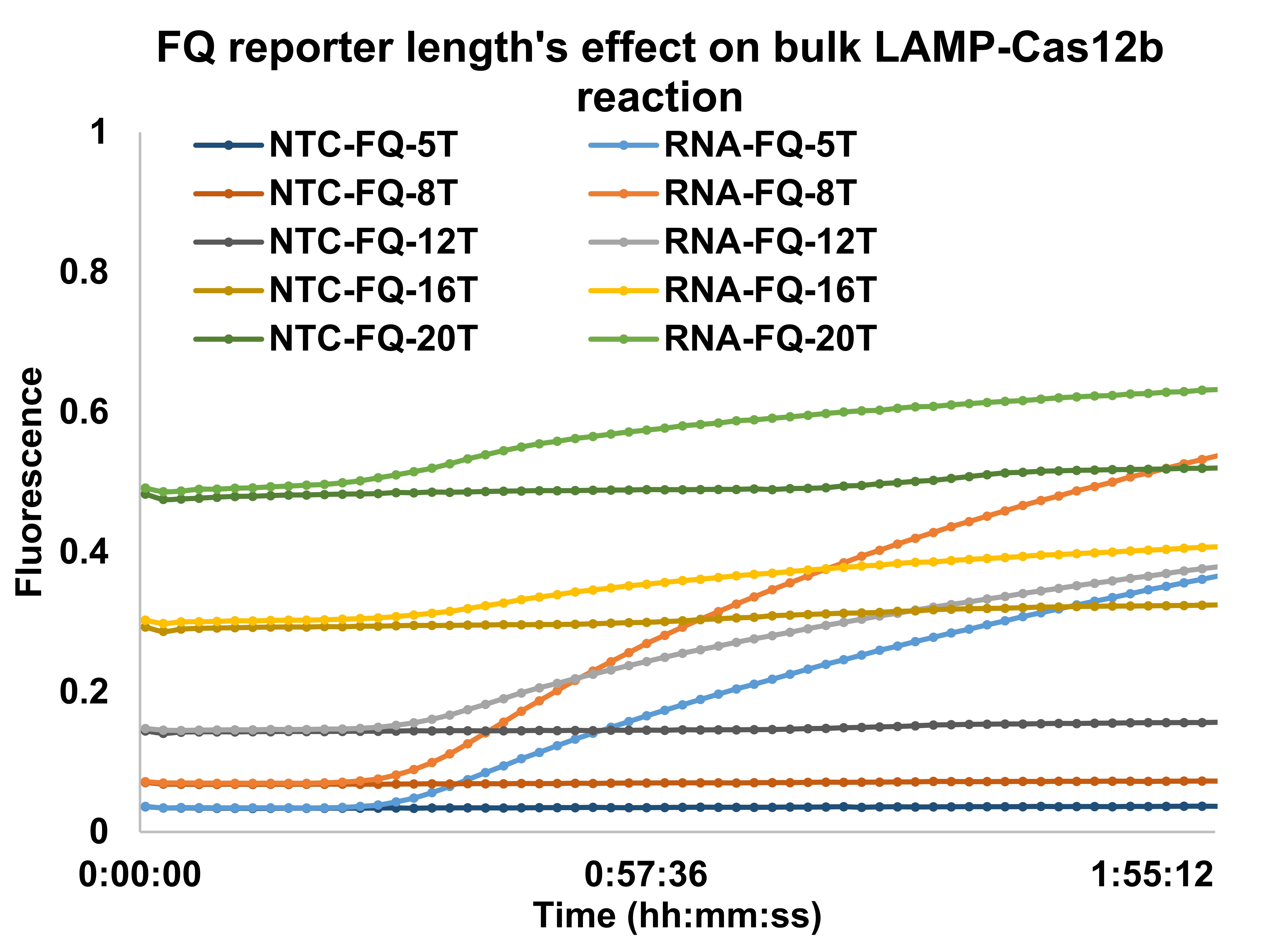

**Supplementary Fig. 2. FQ reporter length's effect on bulk RT-LAMP-Cas12b reaction.** Bulk RT-LAMP-Cas12b reactions with different FQ reporter lengths were monitored at 60 ºC. Fluorescent signals of reactions with target RNA and non-template control were compared.

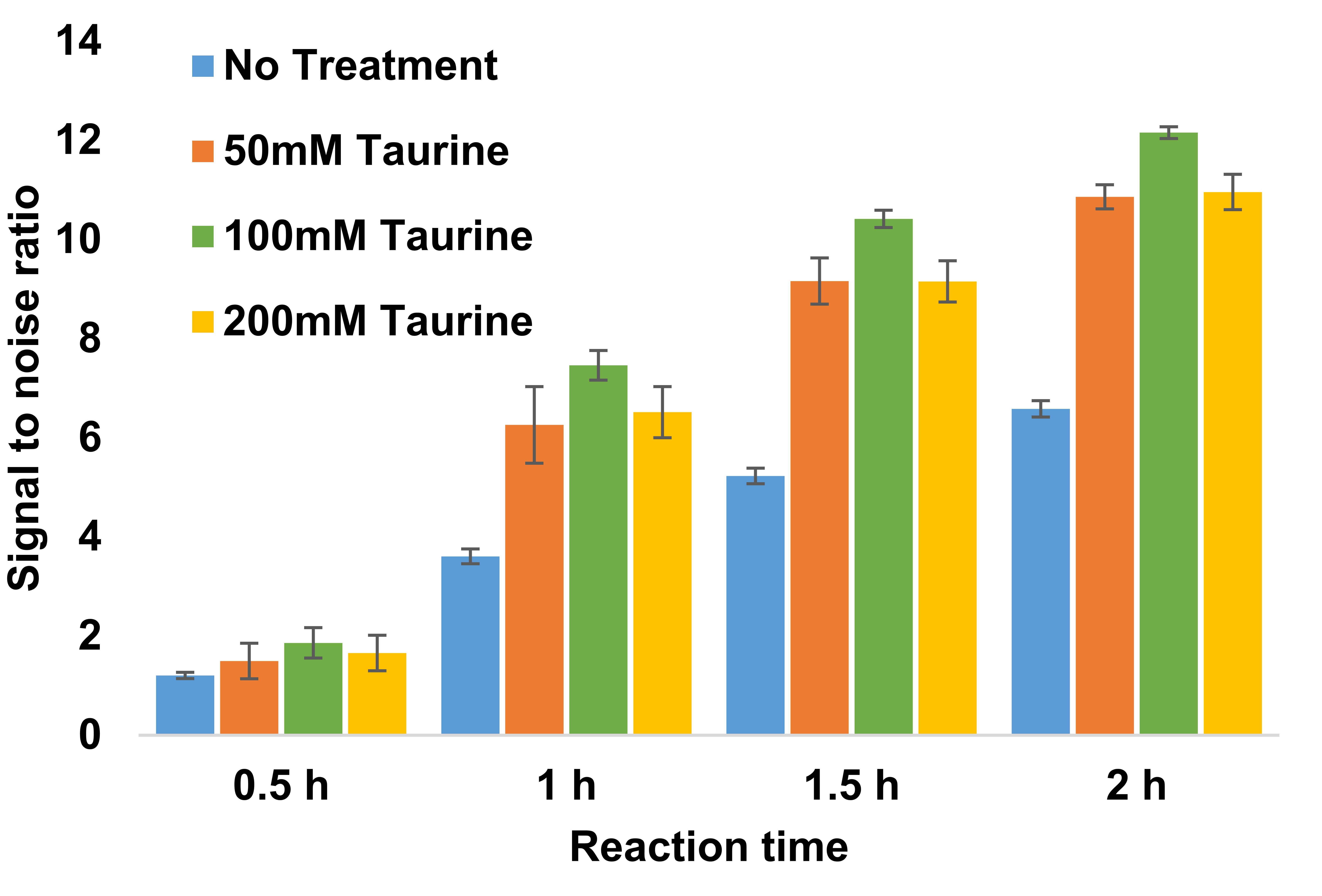

**Supplementary Fig. 3. The effect of taurine on bulk RT-LAMP-Cas12b reaction.** Bulk RT-LAMP-Cas12b reactions with different taurine were monitored at 60 ºC. Fluorescent signals of reactions with target RNA and non-template control were compared and signal-to-noise ratios were calculated as Y-axis. At least three replicates were used for each concentration and error bars indicate the standard deviation of the replicates.

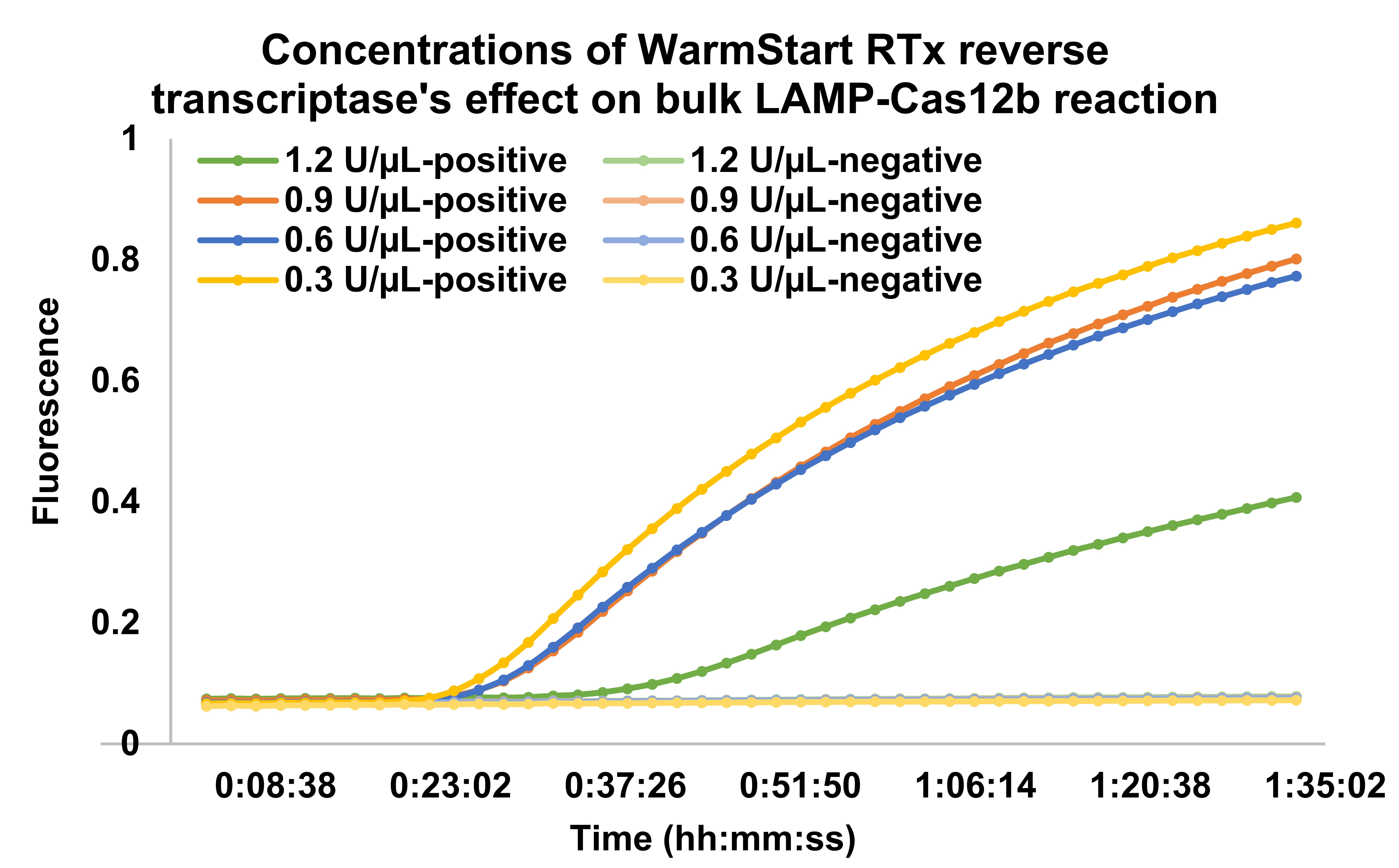

**Supplementary Fig. 4. Concentrations of WarmStart RTx reverse transcriptase's effect on bulk LAMP-Cas12b reaction.** Bulk RT-LAMP-Cas12b reactions with different reverse transcriptase concentrations were monitored at 60 ºC. Fluorescent signals of reactions with target RNA and non-template control were compared.

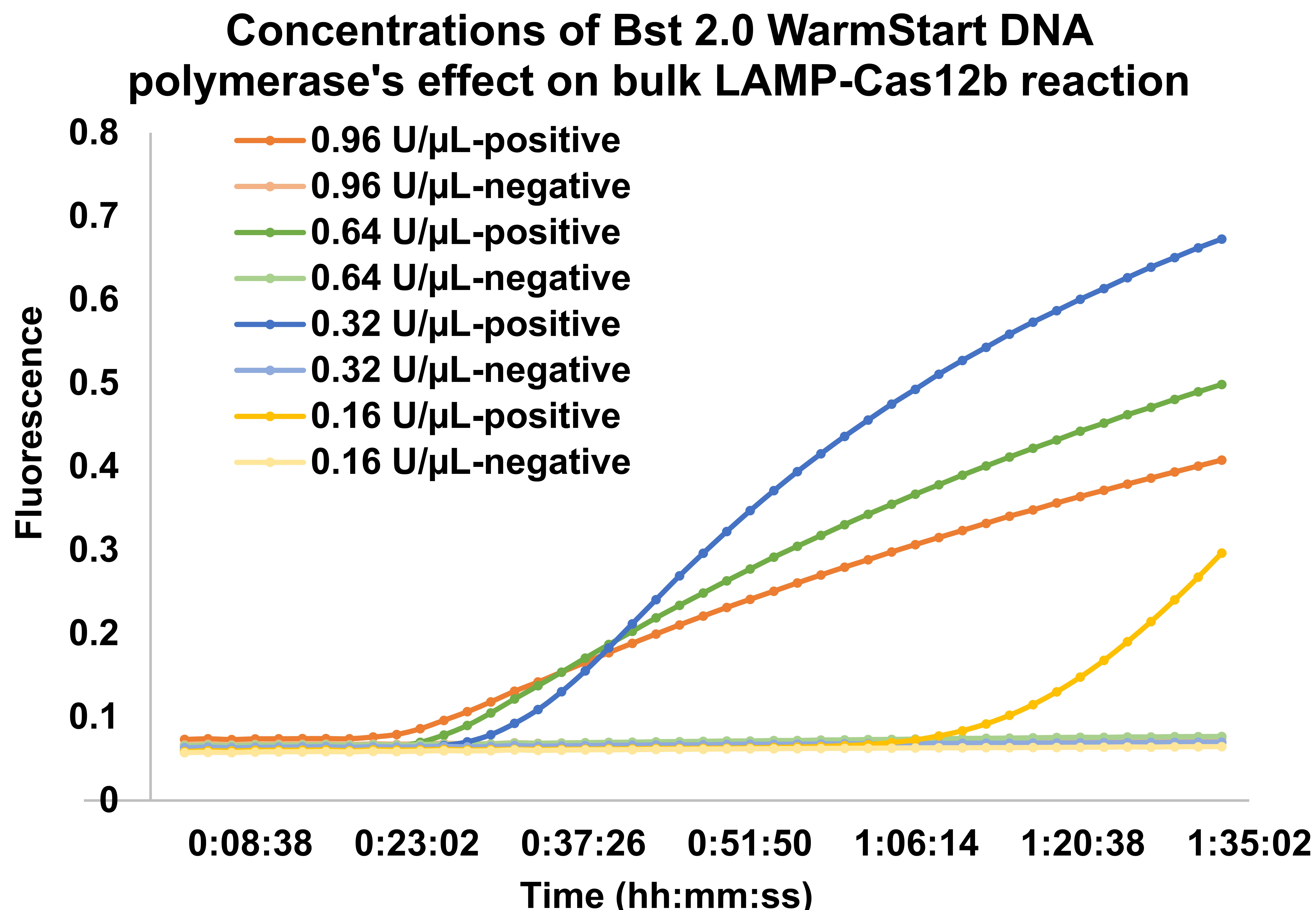

**Supplementary Fig. 5. Concentrations of Bst 2.0 WarmStart DNA polymerase's effect on bulk LAMP-Cas12b reaction.** Bulk RT-LAMP-Cas12b reactions with different polymerase concentrations were monitored at 60 ºC. Fluorescent signals of reactions with target RNA and non-template control were compared.

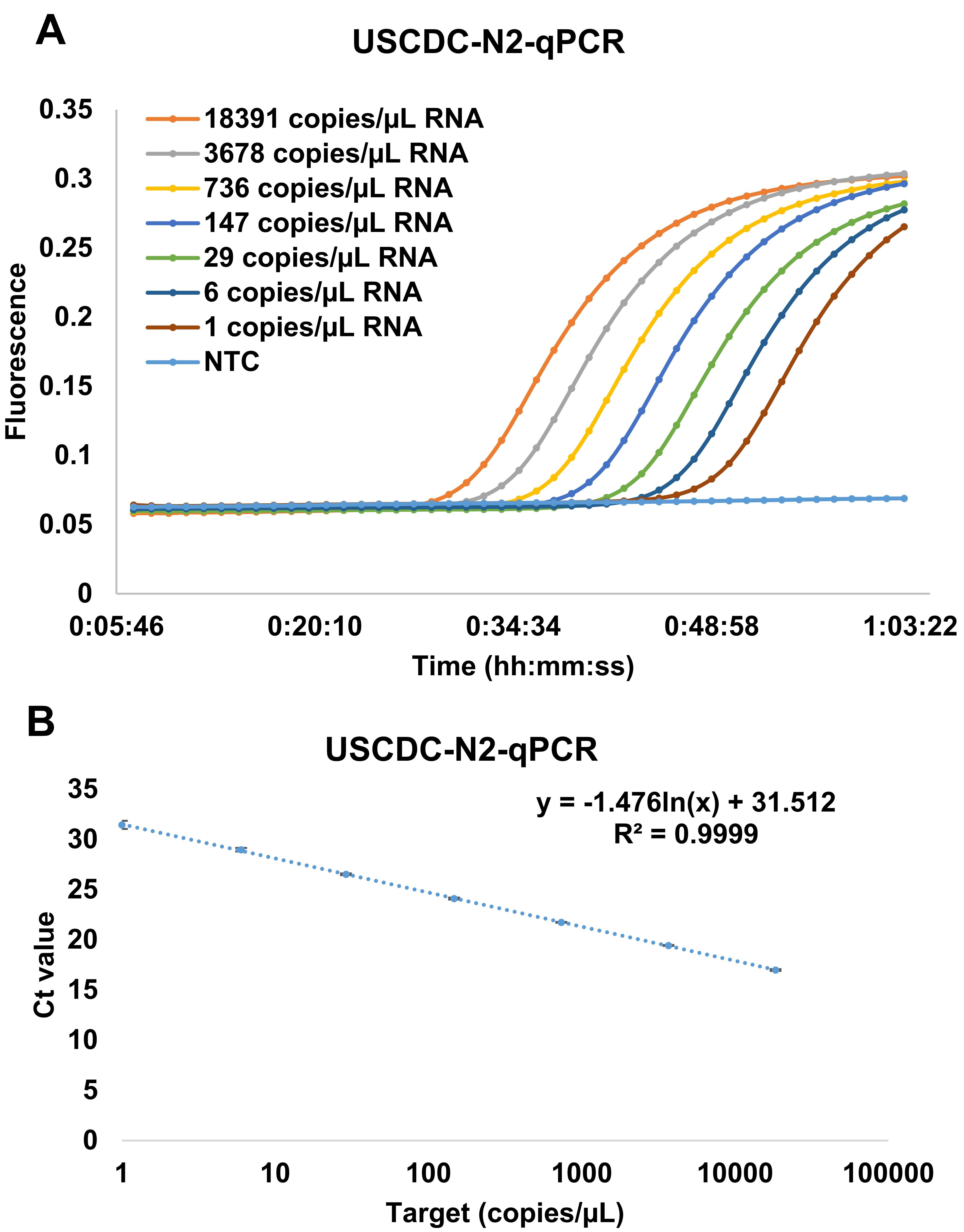

**Supplementary Fig. 6. RT-qPCR assay on different concentrations of SARS-CoV-2 RNA.** (A) Real-time fluorescence curves of RT-qPCR on SARS-CoV-2 RNA. Primers and probes in USCDC N2 assay were used. (B) The standard curve of RT-qPCR to quantify the SARS-CoV-2 RNA. At least three replicates were used for each concentration and error bars indicate the standard deviation of the replicates.

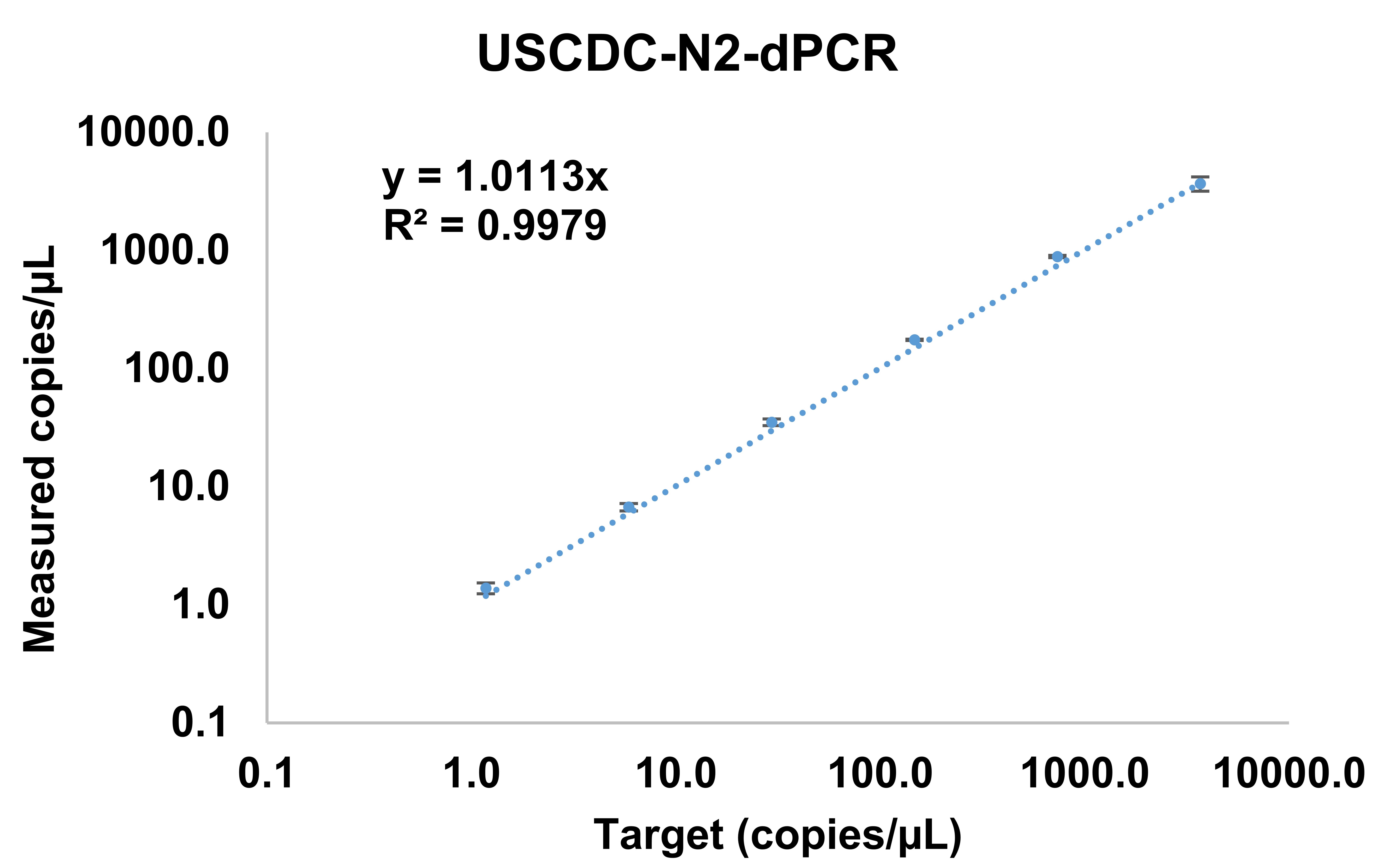

**Supplementary Fig. 7. The correlation between input target SARS-CoV-2 concentration and the measured copy number by RT-dPCR.** Primers and probes in USCDC N2 assay were used.

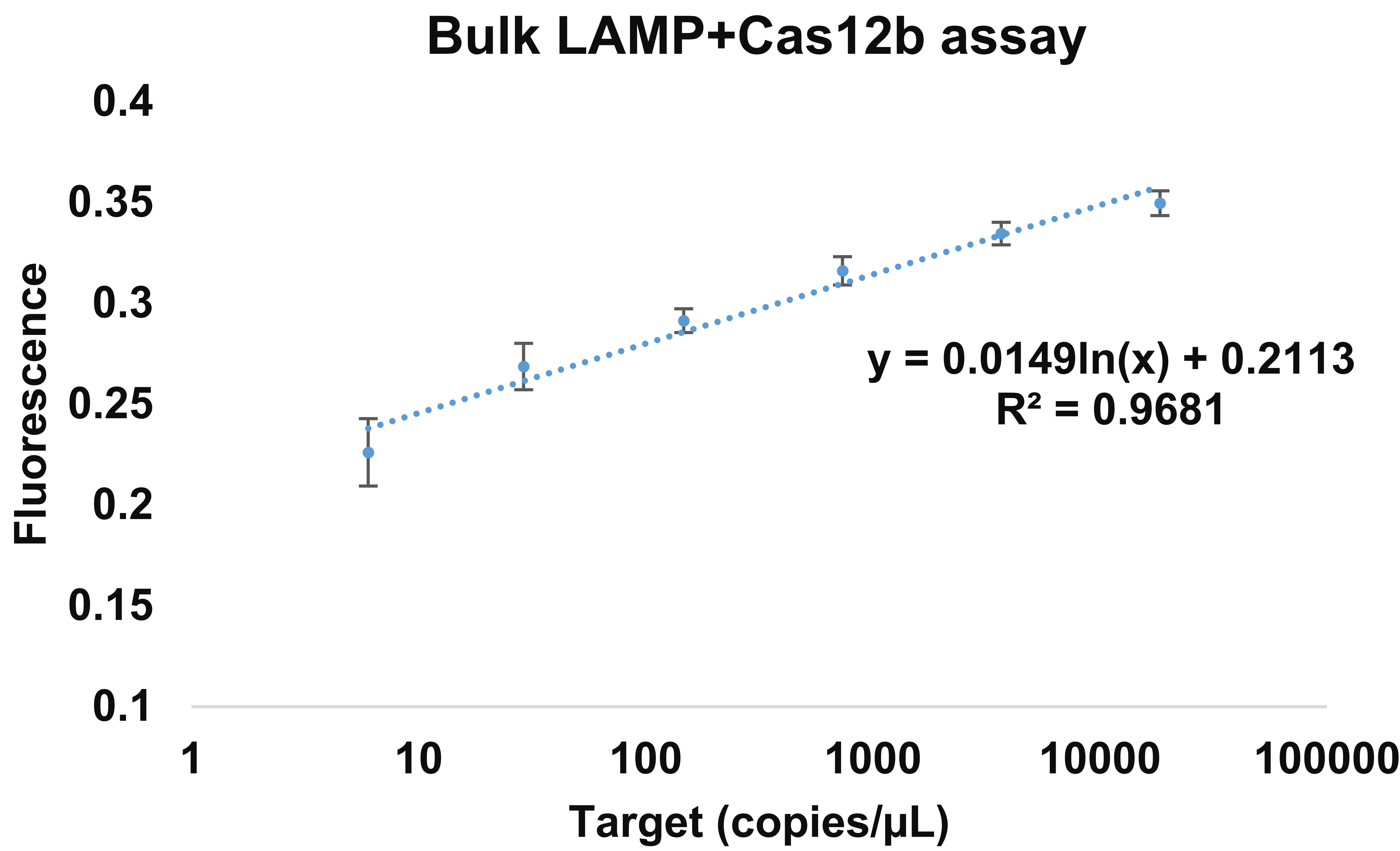

**Supplementary Fig. 8. The standard curve of bulk RT-LAMP-Cas12b to quantify the SARS-CoV-2 RNA.** End-point fluorescent signals after 1 h incubation at 60 ºC were used. At least three replicates were used for each concentration and error bars indicate the standard deviation of the replicates.

| **Name** | **Sequence** | **Application** |
| --- | --- | --- |
| FQ5T | /56-FAM/TTTTT/3IABkFQ/ | FQ reporter |
| FQ8T | /56-FAM/TTTTTTTT/3IABkFQ/ | FQ reporter |
| FQ12T | /56-FAM/TTTTTTTTTTTT/3IABkFQ/ | FQ reporter |
| FQ16T | /56-FAM/TTTTTTTTTTTTTTTT/3IABkFQ/ | FQ reporter |
| FQ20T | /56-FAM/TTTTTTTTTTTTTTTTTTTT/3IABkFQ/ | FQ reporter |
| FQ8A | /56-FAM/AAAAAAAA/3IABkFQ/ | FQ reporter |
| FQ8G | /56-FAM/GGGGGGGG/3IABkFQ/ | FQ reporter |
| FQ8C | /56-FAM/CCCCCCCC/3IABkFQ/ | FQ reporter |
| FQ8AT | /56-FAM/TTATTATT/3IABkFQ/ | FQ reporter |
| N2-LAMP-crRNA11 | GUCUAGAGGACAGAAUUUUUCAACGGGUGUGCCAAUGGCCACUUUCCAGGUGGCAAAGCCCGUUGAGCUUCUCAAAUCUGAGAAGUGGCACCGAAGAACGCUGAAGCGCUG | crRNA for Cas12b^1^ |
| N2-WSLAMP-F3 | GCTGCTGAGGCTTCTAAG | LAMP primers^1^ |
| N2-WSLAMP-B3 | GCGTCAATATGCTTATTCAGC | LAMP primers^1^ |
| N2-WSLAMP-FIP | GCGGCCAATGTTTGTAATCAGTAGACGTGGTCCAGAACAA | LAMP primers^1^ |
| N2-WSLAMP-BIP | TCAGCGTTCTTCGGAATGTCGCTGTGTAGGTCAACCACG | LAMP primers^1^ |
| N2-WSLAMP-LoopF | CCTTGTCTGATTAGTTCCTGGT | LAMP primers^1^ |
| N2-WSLAMP-LoopB | TGGCATGGAAGTCACACC | LAMP primers^1^ |

**Supplementary Table 1. Primers, probes and crRNAs used in this study.**

| **Technology** | **RT-qPCR** | **RT-LAMP-Cas12b bulk reaction** | **RT-dPCR** | **WSRADICA** |
| --- | --- | --- | --- | --- |
| **Result Time** | **1 h** | **1 h** | **3 h** | **1h** |
| **Thermal cycling Requirements** | **Yes** | **No** | **Yes** | **No** |
| **Sensitivity** | **1 copy/µL** | **6 copy/µL** | **1 copy/µL** | **1 copy/µL** |
| **Specificity** | **high** | **high** | **high** | **high** |
| **Quantification** | **Yes** | **No/Bad** | **Yes** | **Yes** |
| **Tolerance to inhibitors** | **No** | **No** | **Yes** | **Yes** |

**Supplementary Table 2. Comparison of WS-RADICA with other viral detection methods.
Supplementary References**

1.	Joung J, Ladha A, Saito M, Kim N-G, Woolley AE, Segel M*, et al.* Detection of SARS-CoV-2 with SHERLOCK One-Pot Testing. *New England Journal of Medicine* 2020.
